# Risk of Hypertensive Disorders of Pregnancy in Patients With Cardiac Disease

**DOI:** 10.1101/2025.01.03.25319981

**Authors:** Maura Jones, Johanna Quist-Nelson, Matthew Fuller, Elizabeth Volz, Sarah Snow, Ashraf S Habib, Jerome Federspiel, Kim Boggess, Marie-Louise Meng

## Abstract

**Background:** Pregnant patients with cardiovascular disease (CVD) face increased risk of preeclampsia and preterm delivery, yet data is limited data regarding degree of risk and impact of hypertensive disorders of pregnancy (HDP) on gestational age at delivery.

**Objectives:** To examine HDP risk and impact on delivery timing in patients with CVD.

**Methods:** This retrospective cohort study included patients >18 years old who delivered between 10/1/2015 and 12/31/2020 using the Premier Healthcare Database. Patients with CVD were divided into six categories: congenital, ischemic, aortic pathology, pulmonary hypertension, cardiomyopathy, and valvular disease. The primary outcome was risk of HDP (gestational hypertension/ preeclampsia). The secondary outcome was gestational age at delivery. Multivariable mixed effects regression models were used to estimate adjusted outcomes, adjusting for CVD subtype, >1 CVD subtype present, demographics, hospital characteristics, and comorbidities.

**Results:** Among 4,606,247 obstetric patients, 20,021 had CVD. The risk of HDP among people with CVD varied by CVD subtype, lowest in those with congenital heart disease (aOR,0.9; 95% CI [0.8, 1.0]) and highest in those with pulmonary hypertension (aOR, 1.5; 95% CI [1.3, 1.8]) and cardiomyopathy (aOR,1.5; 95% CI [1.4,1.6]). Patients with CVD delivered earlier than those without CVD, even in the absence of HDP(36.4–38.0 weeks versus 38.4 weeks). Among those with HDP, patients with severe pre-eclampsia with CVD, delivered earlier than those without CVD (33.1–34.6 weeks versus 35.5 weeks).

**Conclusion:** Risk of HDP and preterm delivery is higher those with CVD, particularly in pulmonary hypertension and cardiomyopathy. Patients with CVD should be advised of their increased risk.

## BACKGROUND

Preeclampsia complicates 2-8% of pregnancies and is a leading cause of maternal morbidity and medically-indicated preterm birth in the United States [1]. Similarly, cardiovascular disease (CVD) is a leading cause of maternal morbidity and mortality, with increasing prevalence, particularly of acquired cardiac disease, in pregnant patients in the United States [2-3].

Among patients with asymptomatic CVD, studies indicate that early-term delivery (delivery between 37-38 weeks of gestation) does not improve maternal cardiovascular or obstetric outcomes but does increase adverse neonatal outcomes when compared with delivery at 39 weeks gestation [4-6]. In line with this, the American College of Obstetricians and Gynecologists (ACOG) recommends delivery at 39 weeks in those with asymptomatic cardiac disease but does not make additional recommendations regarding delivery timing in this patient population, deferring to the managing Pregnancy Heart Team. Conversely, in patients with hypertensive disorders of pregnancy, recommendations regarding delivery timing are well-delineated by ACOG and oftentimes require preterm (34 0/7 weeks) or early-term birth (37 0/7 weeks) [7].

Patients with cardiac disease are at an increased risk of both hypertensive disorders of pregnancy as well as preterm delivery. Preeclampsia, in particular, can precipitate decompensation in previously stable or asymptomatic patients with CVD, increasing the risk of maternal death, heart failure, cesarean delivery, preterm birth, small for gestational age neonates, and perinatal death [8]. However the impact of hypertensive disorders on delivery timing in this population has not been well characterized, nor has delivery timing been examined by specific CVD subtype. [Clarifying these overlapping risk profiles is critical for guiding management and counseling patients about potential complications and delivery decisions.

Given the limited data on delivery timing in patients with CVD who develop hypertensive disorders of pregnancy, our study aimed to describe the risk of hypertensive disorders of pregnancy according to CVD subtype. We also sought to characterize how developing a hypertensive disorder of pregnancy affects delivery timing in patients with CVD in the United States—specifically detailing the gestational week of delivery, as this greatly impacts neonatal morbidity [9,10].

## METHODS

### Study Design and Population

This retrospective cohort study used the Premier Healthcare Database (Premier Inc., Charlotte, North Carolina). Details of the dataset and readmission data have been previously described [11]. The cohort for this study consisted of pregnant patients 18 years of age or older delivering between 10/1/2015 and 12/31/2020, who had both an International Classification of Diseases, Tenth Edition (ICD-10) diagnosis code for delivery after 25 weeks gestation (Z3A.25-Z3A.42) and for cardiac disease (Supplemental Table 1). Included patients with heart disease were divided into six subtypes: congenital, ischemia, arrhythmia, pulmonary hypertension, cardiomyopathy, and valvular disease (Supplemental Table 1). Patients with codes for multiple subtypes of CVD were included in the analyses for each of these CVD categories. This study was deemed exempt from review by the Duke University Health System Institutional Review Board and follows the Strengthening the Reporting of Observational Studies in Epidemiology (STROBE) guidelines [12].

### Exposures, Outcomes, and Covariates

The exposure variable of interest was diagnosis of a hypertensive disorder of pregnancy: gestational hypertension, preeclampsia or preeclampsia with severe features at delivery hospitalization. These categories were considered mutually exclusive and were ascertained by diagnosis codes for gestational hypertension and preeclampsia disorders (Supplemental Table 1). If a patient had multiple hypertensive disorders of pregnancy codes, they were placed in the more severe disease category (i.e., least severe: gestational hypertension, followed by preeclampsia without severe features, followed by preeclampsia with severe features).

The primary outcome was prevalence and risk of hypertensive disorder of pregnancy in patients with CVD, stratified by CVD subtype. The secondary outcome was the gestational age at delivery by CVD subtype and type of hypertensive disorder of pregnancy.

The covariates in the models were selected *a priori* based on known maternal morbidity risk factors, prior literature, and subject-matter expertise: age, payer category (managed care organization, Medicaid, Medicare, and other), and mode of delivery (vaginal, intrapartum cesarean delivery, non-intrapartum cesarean) as well as the presence of more than one category of cardiac disease. The comorbidities present in the expanded obstetric comorbidity index with some adjustments (Supplemental Table 2) were used to define pre-existing comorbid conditions present on admission, however unlike usual practice with the expanded obstetric comorbidity scoring system, we did not include preeclampsia or preeclampsia with severe features as covariates as they are the exposure variables that define the preeclampsia group [13]. Importantly, we treated chronic hypertension as a comorbidity not as a type of HDP.

Some heart failure ICD-10 codes signify chronic disease with an acute heart failure component (Supplemental Table 1). These codes could therefore be used as both inclusion criteria and outcomes; thus, these codes were not included as inclusion criteria in the primary analysis but as outcomes when not present on admission. A planned sensitivity analysis added these acute-on-chronic codes as inclusion criteria regardless of present on admission status.

### Statistical Analysis

Demographic and clinical characteristics of the cohort are reported, stratified by subtype of CVD. Descriptive statistics were used to examine the study population with categorical variables reported as counts and frequencies, and continuous data reported as median with interquartile range. Associations between CVD categories, hypertensive disorders of pregnancy, and gestational age at delivery were assessed using a multivariable mixed linear regression model. This model included fixed effects for patient and hospital characteristics and a random intercept for each hospital to account for the clustering of patients within centers. The least-squares mean gestational age at delivery was estimated using this model and is reported with an associated 95% confidence interval. A multivariable logistic regression model was used to compare the association between the CVD subtype and the development of hypertensive disorders of pregnancy. This regression model included fixed effects for patient and hospital characteristics as well as a random intercept for hospitals. Association between the CVD subtype and the development of hypertensive disorders of pregnancy are reported as odds ratios with 95% confidence intervals.

Statistical analyses were performed in the SAS System, Version 9.4 (SAS Institute, Cary, North Carolina). A two-sided p-value < 0.05 was pre-specified as statistically significant.

## RESULTS

Of 4,606,247 deliveries, 20,021 (0.43%) deliveries had a diagnosis of CVD present on admission. Baseline demographic characteristics and comorbidities are described in Table 1. The median age among patients with CVD was 31 years [Quartile 1-Quartile 3 (Q1-Q3): 25-34] and 29 years [Q1-Q3: 25-33] in those without CVD. Most patients with CVD delivered in urban, large, or teaching hospitals. Patients with CVD were more likely to have chronic hypertension; ranging from 6.7% in those with congenital heart disease to 30.5% in those with cardiomyopathy, compared to 3.4% in those without CVD.

**Table 1.**
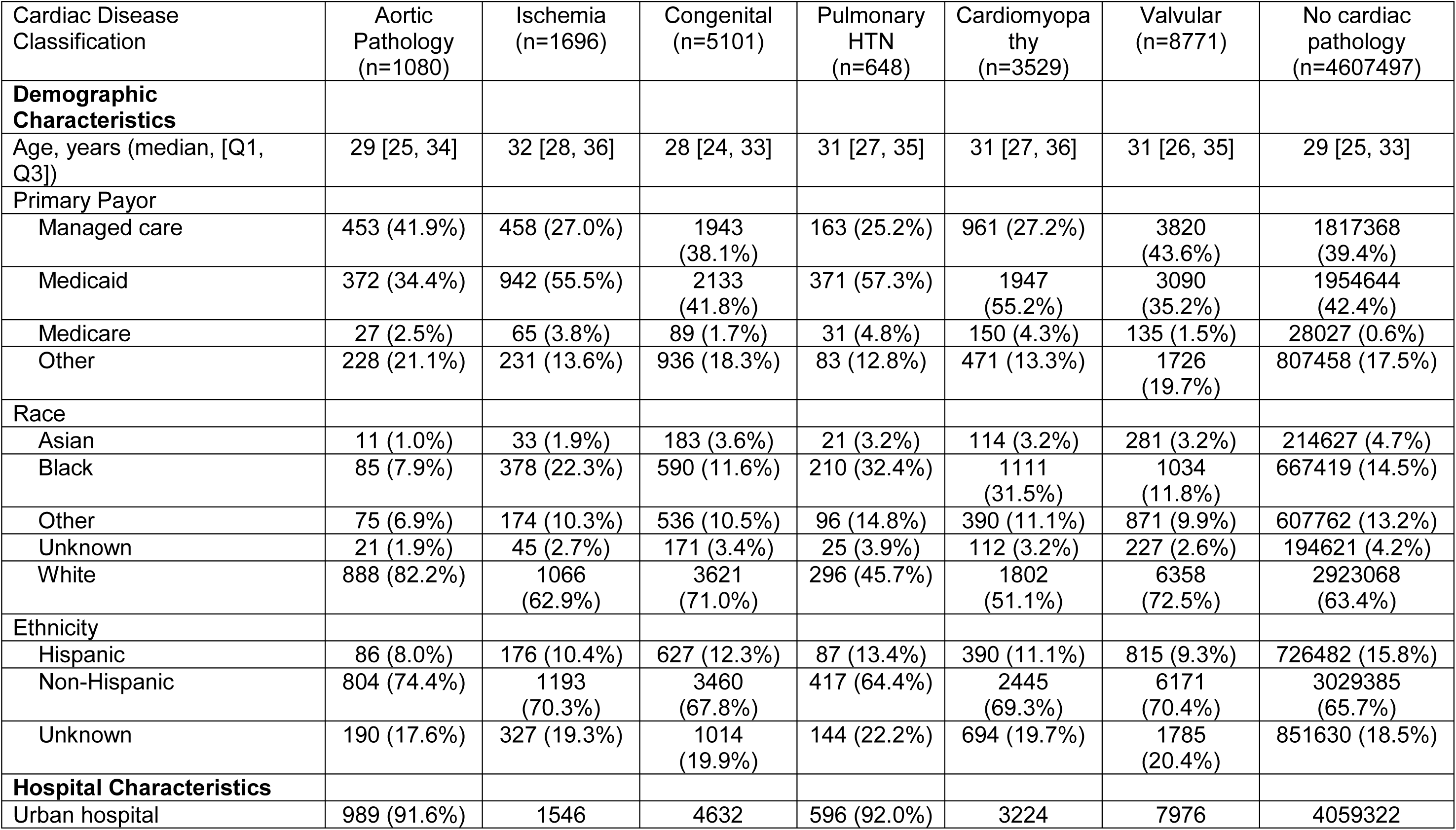
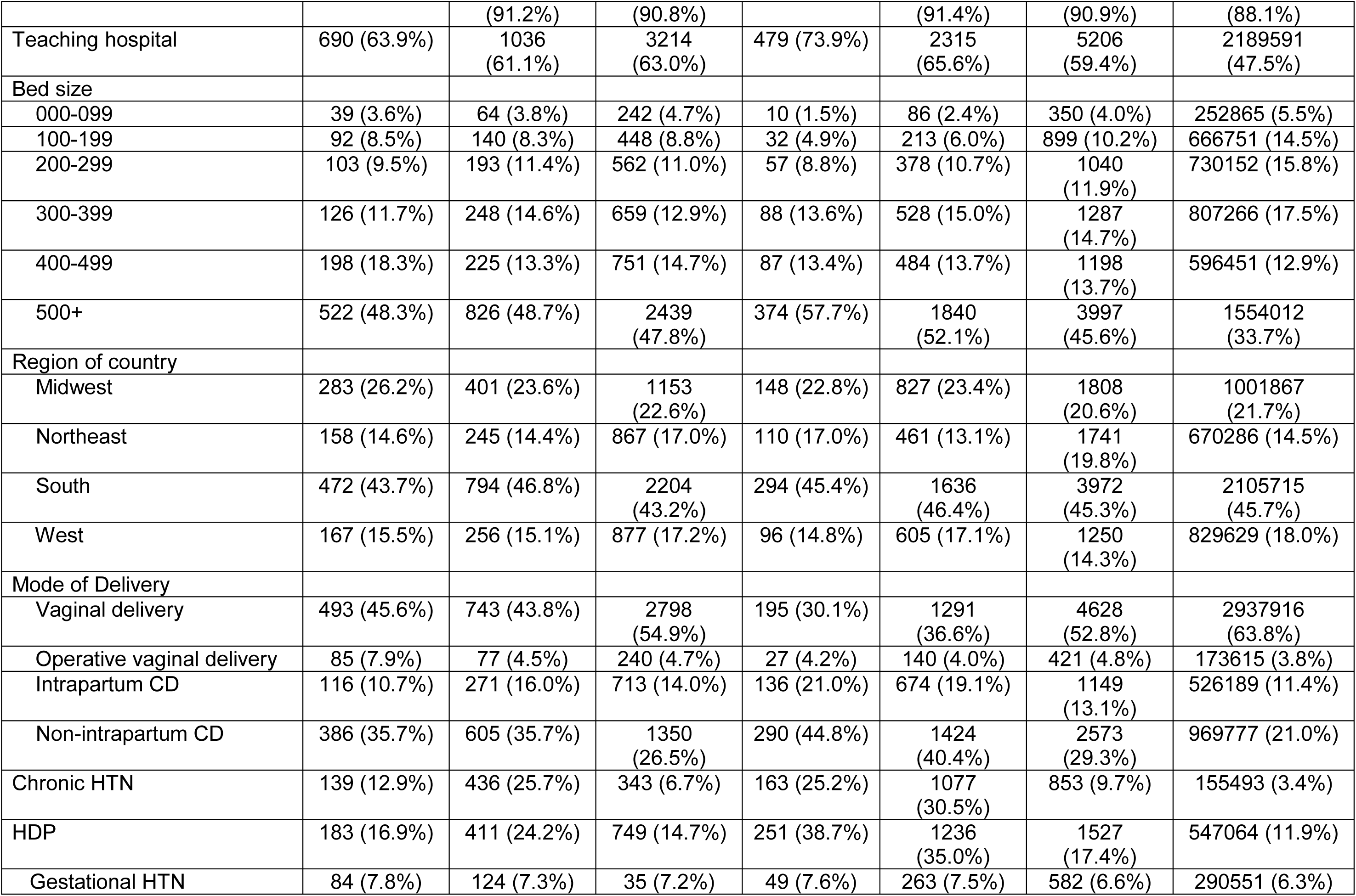
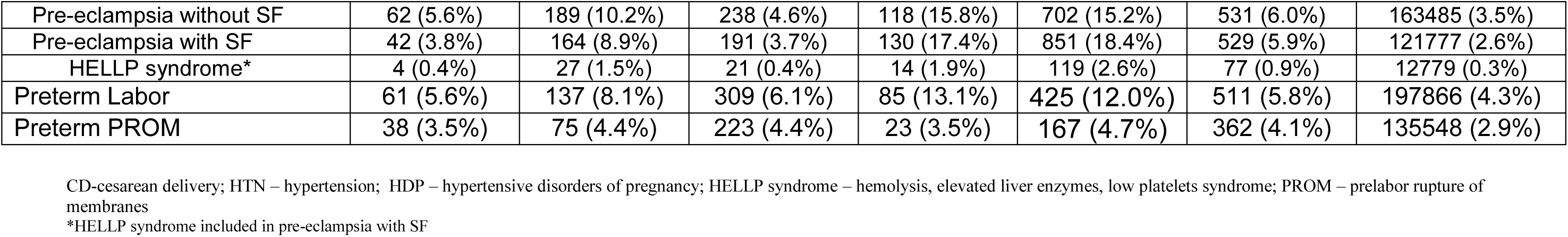
Baseline demographic characteristics of cohort.

### Prevalence and risk of hypertensive disorders of pregnancy

The prevalence of hypertensive disorders of pregnancy in patients without CVD was 11.9%. The rate of hypertensive disorders of pregnancy among people with CVD varied by CVD subtype, with the lowest rate in patients with congenital CVD at 14% and the highest in those with cardiomyopathy and pulmonary hypertension at 38.7% and 35.0%, respectively (Table 1). When controlling for age, payor, race, ethnicity, hospital type, hospital location, hospital size, and comorbidities (including chronic hypertension), the risk of a hypertensive disorder of pregnancy was significantly higher in those with pulmonary hypertension (OR 1.6 [95% CI: 1.3 - 1.8]) and cardiomyopathy (OR 1.5, [CI: 1.4-1.6]) compared to patients without CVD or those particular CVD subtype diagnoses. (Table 2).

**Table 2.** Adjusted Risk of Hypertensive disorder of pregnancy by cardiac disease subtype.

| **Cardiac Disease subtype** | **Risk of HDP\* [OR (95% CI)]** | **p-value** |
| --- | --- | --- |
| Aortic pathology | 0.98 (0.83, 1.17) | 0.851 |
| Ischemia | 0.96 (0.85, 1.09) | 0.553 |
| Congenital | 0.90 (0.82, 0.97) | 0.009 |
| Pulmonary hypertension | 1.52 (1.27, 1.83) | <0.001 |
| Cardiomyopathy | 1.45 (1.34, 1.57) | <0.001 |
| Valvular disease | 1.06 (1.00, 1.12) | 0.073 |

*\*Reference group for each subtype are those without that CVD subtype ( i.e. series of multiple regressions with those without that CVD subtype diagnosis code as referent group)*
HDP – hypertensive disorders of pregnancy

### Delivery Timing

Delivery timing in this cohort is described in Figure 1 and is presented as least-square mean gestational ages reported with an associated 95% confidence interval. On average, patients with CVD delivered at earlier gestational ages across the six subtypes (least squares means ranging from 36.4 – 38.0 weeks) than those without CVD (38.4 weeks), even in the absence of concomitant hypertensive disorders of pregnancy. Patients with hypertensive disorders of pregnancy and CVD, across all subtypes, delivered between 33-37 weeks (least squares means ranging from 33.1 – 37.8 weeks) while patients with hypertensive disorders of pregnancy without CVD delivered at 37 weeks (37.2 weeks). When examining by hypertensive disorder subtype, patients with gestational hypertension plus CVD, across all subtypes, delivered at 36-37 weeks (least squares means ranging from 36.2 – 37.8 weeks) vs at 38 weeks (38.1 weeks) in patients with gestational hypertension without CVD. Patients with pre-eclampsia without severe features (SF) plus CVD, across all subtypes, delivered at 33-36 weeks (least squares means ranging from 33.1 - 36.4 weeks) vs 37 weeks (36.9 weeks) in patients with pre-eclampsia without SF without CVD. Patients with pre-eclampsia with SF plus CVD, across all subtypes, delivered at 33-34 weeks (least squares means ranging from 33.1 – 34.6 weeks) vs 35 weeks (35.5 weeks) in patients with pre-eclampsia with SF without CVD.

**Figure 1.**
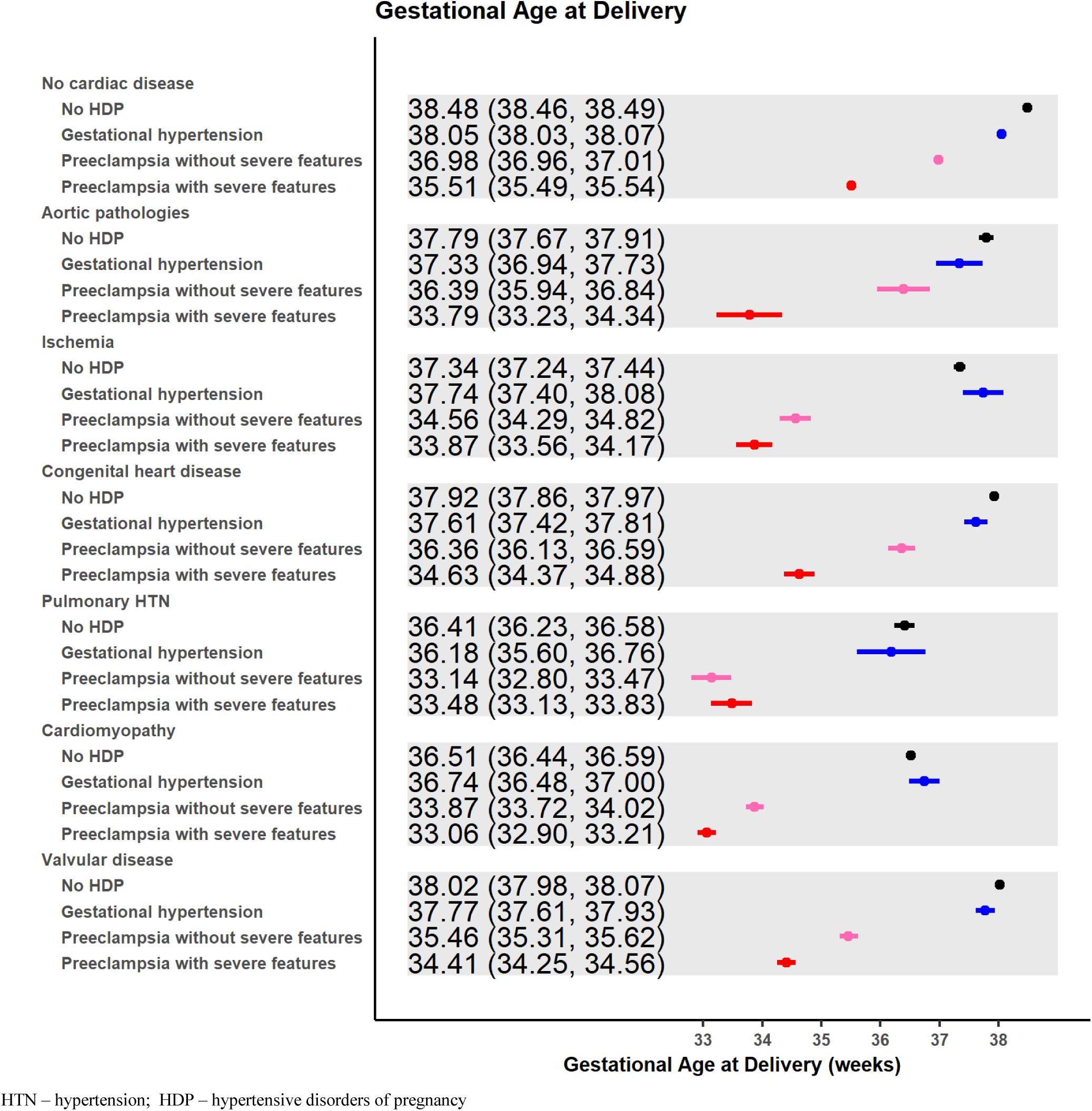
Delivery timing by gestational age and hypertensive disorder of pregnancy in those with cardiac disease, by subtype.

Across all CVD subtypes, those diagnosed with pre-eclampsia with SF delivered 3-4 weeks earlier than those within their same CVD subtype who did not develop hypertensive disorders of pregnancy (Table 3). Patients with pulmonary hypertension or cardiomyopathy delivered earliest regardless of hypertensive disorder of pregnancy diagnosis. Patients with pulmonary hypertension without hypertensive disorder of pregnancy delivered at 36 weeks (36.4 weeks) however if diagnosed with preeclampsia with SF, the mean gestational age at delivery was 33 weeks (33.4 weeks). Similarly, those with cardiomyopathy delivered at 36 weeks (36.5 weeks) if they did not develop hypertensive disorder of pregnancy vs 33 weeks (33.1 weeks) if they developed preeclampsia with severe features (Figure 1).

**Table 3.** Risk of Hypertensive disorder of pregnancy by cardiac disease subtype and hypertensive disorder subtype.

| **Cardiac Disease subtype** | **Least squares mean delivery time (95% CI), weeks** | **Difference (No HDP – HDP)** | **p-value** |
| --- | --- | --- | --- |
| No cardiac disease |  |  |  |
| No HDP | 38.48 (38.46, 38.49) | (reference) |  |
| Gestational HTN | 38.05 (38.03, 38.07) | 0.43 (0.42, 0.44) | <0.001 |
| Preeclampsia | 36.98 (36.96, 37.01) | 1.49 (1.48, 1.50) | <0.001 |
| Preeclampsia with SF | 35.51 (35.49, 35.54) | 2.96 (2.95, 2.97) | <0.001 |
| Aortic pathologies |  |  |  |
| No HDP | 37.79 (37.67, 37.91) | (reference) |  |
| Gestational HTN | 37.33 (36.94, 37.73) | 0.45 (0.05, 0.86) | 0.030 |
| Preeclampsia | 36.39 (35.94, 36.84) | 1.40 (0.94, 1.86) | <0.001 |
| Preeclampsia with SF | 33.79 (33.23, 34.34) | 4.00 (3.43, 4.57) | <0.001 |
| Ischemia |  |  |  |
| No HDP | 37.34 (37.24, 37.44) | (reference) |  |
| Gestational HTN | 37.74 (37.40, 38.08) | -0.40 (-0.75, -0.04) | 0.028 |
| Preeclampsia | 34.56 (34.29, 34.82) | 2.78 (2.50, 3.07) | <0.001 |
| Preeclampsia with SF | 33.87 (33.56, 34.17) | 3.47 (3.15, 3.79) | <0.001 |
| Congenital disorders |  |  |  |
| No HDP | 37.92 (37.86, 37.97) | (reference) |  |
| Gestational HTN | 37.61 (37.42, 37.81) | 0.30 (0.10, 0.50) | 0.003 |
| Preeclampsia | 36.36 (36.13, 36.59) | 1.56 (1.33, 1.79) | <0.001 |
| Preeclampsia with SF | 34.63 (34.37, 34.88) | 3.29 (3.03, 3.55) | <0.001 |
| Pulmonary HTN |  |  |  |
| No HDP | 36.41 (36.23, 36.58) | (reference) |  |
| Gestational HTN | 36.18 (35.60, 36.76) | 0.23 (-0.38, 0.83) | 0.462 |
| Preeclampsia | 33.14 (32.80, 33.47) | 3.27 (2.89, 3.65) | <0.001 |
| Preeclampsia with SF | 33.48 (33.13, 33.83) | 2.93 (2.54, 3.31) | <0.001 |
| Cardiomyopathy |  |  |  |
| No HDP | 36.51 (36.44, 36.59) | (reference) |  |
| Gestational HTN | 36.74 (36.48, 37.00) | -0.23 (-0.50, 0.04) | 0.094 |
| Preeclampsia | 33.87 (33.72, 34.02) | 2.64 (2.48, 2.81) | <0.001 |
| Preeclampsia with SF | 33.06 (32.90, 33.21) | 3.46 (3.29, 3.63) | <0.001 |
| Valvular disease |  |  |  |
| No HDP | 38.02 (37.98, 38.07) | (reference) |  |
| Gestational HTN | 37.77 (37.61, 37.93) | 0.26 (0.10, 0.42) | 0.002 |
| Preeclampsia | 35.46 (35.31, 35.62) | 2.56 (2.40, 2.72) | <0.001 |
| Preeclampsia with SF | 34.41 (34.25, 34.56) | 3.62 (3.46, 3.78) | <0.001 |

HDP – hypertensive disorders of pregnancy, HTN - Hypertension

## DISCUSSION

This large retrospective study of obstetric patients found that patients with pulmonary hypertension and cardiomyopathy have increased risk of hypertensive disorders of pregnancy compared with obstetric patients without CVD or with other subtypes of CVD. Patients with CVD delivered earlier than patients without CVD, regardless of concomitant hypertensive disorders of pregnancy. Patients with CVD and hypertensive disorders of pregnancy delivered two to three weeks earlier than patients without CVD and hypertensive disorders of pregnancy, with those with pre-eclampsia with SF delivering earliest.

As previously noted, the prevalence of CVD is rising among patients of childbearing age and remains a leading cause of maternal morbidity and mortality. Early, individualized counseling regarding maternal and fetal risks is therefore essential, and low-dose aspirin should be recommended for preeclampsia prevention. [14]. These results indicate that pregnant patients with CVD are at significantly increased risk of preterm birth, especially if diagnosed with a hypertensive disorder of pregnancy. While ACOG recommends patients with asymptomatic CVD deliver at 39 weeks, these data suggest that patients with cardiac disease are routinely delivered early term, even without concomitant hypertensive disorders of pregnancy [6]. This trend may reflect the presence of maternal symptoms or fetal complications that necessitate earlier delivery in this high-risk population.

In addition, ACOG has identical delivery timing recommendations for patients with preeclampsia without SF and patients with gestational hypertension of 37 weeks, yet in our study, patients with preeclampsia without SF delivered, on average, earlier than those with gestational hypertension, across all cardiac disease subtypes [1]. These findings suggest that within the United States (USA) clinicians may be unlikely to expectantly manage patients with preeclampsia without SF in the context of CVD until recommended delivery timing, or that patients with CVD who have preeclampsia without SF develop indications for earlier delivery.

### Comparison to other studies

Our findings should be considered in the context of other published data. The primarily European study *Hypertensive disorders of pregnant women with heart disease: the ESC EORP ROPAC Registry,* included 5,700 pregnancies among people with CVD and demonstrated a prevalence of 10.3% of hypertensive disorders of pregnancy. They similarly noted an increased prevalence of hypertensive disorders of pregnancy in those with cardiomyopathy (7.1%) and pulmonary hypertension (11.1%). In our study, which is limited to the USA, the prevalence of hypertensive disorders of pregnancy was 11.2%, but significantly higher among similar CVD subtypes when compared to *the ROPAC Registry*, 38.7% in those with cardiomyopathy and 35.0% in those with pulmonary hypertension. An important difference in the *ROPAC Registry* data is that they include chronic hypertension in their definition of hypertensive disorders of pregnancy, with an adjusted prevalence of 6% once chronic hypertension is removed. Conversely, our analysis considered pre-existing chronic hypertension as a maternal comorbidity, and thus, we were able to adjust for its effect on the prevalence of hypertensive disorders of pregnancy.

Our findings are consistent with a 2017 US-based study examining trends and in-hospital outcomes of women with heart disease utilizing the National Inpatient Sample. Compared to the *ROPAC Registry*, they too reported an increased prevalence of preeclampsia, in patients with CVD (11.4%) most notable in cardiomyopathy (25.6%) and pulmonary hypertension (22.3%) [3]. These data highlight a notably increased risk of hypertensive disorders of pregnancy that exists in an American cohort. The higher rate of hypertensive disorders of pregnancy observed in the Premier dataset compared to the ROPAC registry may reflect broader inclusion criteria and reliance on administrative coding practices in a U.S.-based dataset versus the more focused recruitment of severe cardiac cases in a European registry. Additionally, regional differences in population characteristics, healthcare systems, and clinical practices further contribute to this discrepancy.

The *ROPAC Registry* similarly noted that those with hypertensive disorders of pregnancy had higher rates of preterm delivery. Delivery timing in the *ROPAC Registry* study was stratified as early onset (<34 weeks) or late onset (>34 weeks) without further clarification of gestational age at delivery [8]. It is known that a continuum of outcomes exists regarding neonatal morbidity and mortality, with each additional week conferring increased benefit [9-10]. Therefore, we sought to explain further differences in delivery timing, specifically by weeks of gestational age, observed in patients with known CVD and hypertensive disorders of pregnancy versus those without CVD. In addition, while patients with CVD have traditionally been analyzed as a single group, we sought to clarify the risks of hypertensive disorders of pregnancy and preterm birth by examining six distinct CVD subtypes. This approach builds on the framework of the ROPAC Registry but utilizes a more generalizable American cohort.

### Strengths

Our study has several strengths, Our sample size, four times that of ROPAC, allowed us to have power to detect a difference in delivery timing in patients with hypertensive disorders of pregnancy versus those without hypertensive disorders of pregnancy in the six most common CVD subtypes. Though we noted increased prevalence of hypertensive disorders of pregnancy among specific CVD subtype, the overall prevalence of pre-eclampsia in our study was 5.8%, compared to the general population estimate of 2-8% for preeclampsia, indicating the Premier dataset is representative of the US population and, therefore, results are likely generalizable to high-income nations.

### Limitations

Despite these strengths, limitations exist. Administrative database analyses are limited in that chart reviews cannot be performed to adjudicate diagnoses or examine other factors that may have influenced delivery timing, such as timing of hypertensive disorders of pregnancy diagnosis, severity of CVD, worsening of medical comorbidities, cardiac decompensation, or fetal status. While we suspect patients with underlying CVD who develop hypertensive disorders of pregnancy are deferred expectant management and thus deliver earlier, there may be differences in the timing of onset of hypertensive disease in those with CVD versus those without, that we are unable to ascertain.

### Conclusion

In conclusion, our study highlights the significant impact of hypertensive disorders of pregnancy on the timing of delivery in patients with CVD. Patient counseling should ensure that women with CVD are fully informed about their heightened risk of hypertensive disorders of pregnancy and preterm birth.

Future studies should similarly attempt to disaggregate data by CVD subtype, include the timing of hypertensive disorder of pregnancy diagnosis, and focus on morbidity and mortality during delivery hospitalization for the first year postpartum to further inform patient counseling and offer a more nuanced and comprehensive understanding of how to optimize care.

## Data Availability

Data for this study were derived from the Premier Healthcare Database (Premier, Inc., Charlotte, NC), a large, U.S. hospital-based administrative database. The Premier Healthcare Database is proprietary and subject to licensing and confidentiality agreements; therefore, the data are not publicly available. Interested researchers may apply for access to these data by contacting Premier, Inc. directly. The authors do not have the authority to share these data.

## Acknowledgments

None

## REFERENCES

1. Gestational Hypertension and Preeclampsia: ACOG Practice Bulletin Summary, Number 222. Obstet Gynecol. 2020 Jun;135(6):1492-1495. doi: 10.1097/AOG.0000000000003892. PMID: 32443077.

2. Petersen EE, Davis NL, Goodman D, et al. *Vital Signs:* Pregnancy-Related Deaths, United States, 2011–2015, and Strategies for Prevention, 13 States, 2013–2017. MMWR Morb Mortal Wkly Rep 2019;68:423–429. DOI: 10.15585/mmwr.mm6818e1

3. Lima FV, Yang J, Xu J, Stergiopoulos K. National Trends and In-Hospital Outcomes in Pregnant Women With Heart Disease in the United States. Am J Cardiol. 2017 May 15;119(10):1694–1700. doi: 10.1016/j.amjcard.2017.02.003. Epub 2017 Feb 28. PMID: 28343597.

4. Rouse CE, Easter SR, Duarte VE, Drakely S, Wu FM, Valente AM, Economy KE. Timing of Delivery in Women with Cardiac Disease. Am J Perinatol. 2022 Aug;39(11):1196–1203. doi: 10.1055/s-0040-1721716. Epub 2020 Dec 22. PMID: 33352586.

5. Mok T, Woods A, Small A, Canobbio MM, Tandel MD, Kwan L, Lluri G, Reardon L, Aboulhosn J, Lin J, Afshar Y. Delivery Timing and Associated Outcomes in Pregnancies With Maternal Congenital Heart Disease at Term. J Am Heart Assoc. 2022 Aug 16;11(16):e025791. doi: 10.1161/JAHA.122.025791. Epub 2022 Aug 9. PMID: 35943056; PMCID: PMC9496287.

6. American College of Obstetricians and Gynecologists’ Presidential Task Force on Pregnancy and Heart Disease and Committee on Practice Bulletins—Obstetrics. ACOG Practice Bulletin No. 212: Pregnancy and Heart Disease. Obstet Gynecol. 2019 May;133(5):e320-e356. doi: 10.1097/AOG.0000000000003243. PMID: 31022123.

7. American College of Obstetricians and Gynecologists’ Committee on Obstetric Practice, Society for Maternal-Fetal Medicine. Medically Indicated Late-Preterm and Early-Term Deliveries: ACOG Committee Opinion, Number 831. Obstet Gynecol. 2021 Jul 1;138(1):e35-e39. doi: 10.1097/AOG.0000000000004447. PMID: 34259491.

8. Ramlakhan KP, Malhamé I, Marelli A, Rutz T, Goland S, Franx A, Sliwa K, Elkayam U, Johnson MR, Hall R, Cornette J, Roos-Hesselink JW. Hypertensive disorders of pregnant women with heart disease: the ESC EORP ROPAC Registry. Eur Heart J. 2022 Oct 11;43(38):3749–3761. doi: 10.1093/eurheartj/ehac308. PMID: 35727736; PMCID: PMC9840477

9. Ancel P, Goffinet F, and the EPIPAGE-2 Writing Group. Survival and Morbidity of Preterm Children Born at 22 Through 34 Weeks’ Gestation in France in 2011: Results of the EPIPAGE-2 Cohort Study. JAMA Pediatr. 2015;169(3):230–238. doi:10.1001/jamapediatrics.2014.3351

10. Manuck TA, Rice MM, Bailit JL, Grobman WA, Reddy UM, Wapner RJ, Thorp JM, Caritis SN, Prasad M, Tita AT, Saade GR, Sorokin Y, Rouse DJ, Blackwell SC, Tolosa JE; Eunice Kennedy Shriver National Institute of Child Health and Human Development Maternal-Fetal Medicine Units Network. Preterm neonatal morbidity and mortality by gestational age: a contemporary cohort. Am J Obstet Gynecol. 2016 Jul;215(1):103.e1-103.e14. doi: 10.1016/j.ajog.2016.01.004. Epub 2016 Jan 7. PMID: 26772790; PMCID: PMC4921282.

11. Meng ML, Fuller M, Federspiel JJ, et al. Maternal morbidity according to mode of delivery among pregnant patients with pulmonary hypertension. Anesth Analg. Published online May 16, 2023. 10.1213/ANE.0000000000006523

12. von Elm E, Altman DG, Egger M, et al. The Strengthening the Reporting of Observational Studies in Epidemiology (STROBE) statement: guidelines for reporting observational studies. Lancet. 2007;370:1453–1457.

13. Leonard SA, Kennedy CJ, Carmichael SL, Lyell DJ, Main EK. An expanded obstetric comorbidity scoring system for predicting severe maternal morbidity. Obstet Gynecol. 2020;136: 440–449.

14. ACOG Committee Opinion No. 743: Low-Dose Aspirin Use During Pregnancy. Obstet Gynecol. 2018 Jul;132(1):e44-e52. doi: 10.1097/AOG.0000000000002708. PMID: 29939940.

